# The impact of high dose oral cotrimoxazole in patients with COVID-19 with hypoxic respiratory failure requiring non-invasive ventilation: A Case Control Study

**DOI:** 10.1101/2021.01.14.21249803

**Authors:** Saurabh Singh, Thomas John, Prashant Kumar, Syed Rehan Quadery

**Author notes:** Corresponding Author: Saurabh Singh, Assistant Professor, Department of Respiratory Medicine, IQ City Medical College Hospital, Durgapur, West Bengal, India. Declaration of conflict of interest. None declared. Author approval: All authors have seen and approved the manuscript. Ethics: Ethical approval granted by the IQ City Medical College Hospital Institutional Ethics Committee. Patient consent: No patient consent required as all data has been anonymized and no patient identifiable data included.

## Abstract

**Background:** COVID-19 can be fatal in a significant proportion of people who develop critical illness, resulting in hypoxic respiratory failure secondary to Acute Respiratory Distress Syndrome (ARDS) which is thought to be mediated by a cytokine storm syndrome. Steroids have been shown to be of some benefit, but the mainstay of treatment remains supportive.

**Methods:** The data was collected retrospectively from consecutive, newly diagnosed patients presenting to the critical care facility of I Q City Medical College Hospital, Durgapur, India between June and November 2020 with critical COVID-19 on non-invasive ventilation treated with high dose oral cotrimoxazole (CTX) in addition to standard therapy (ST) and compared with patients with critical COVID-19 receiving standard therapy alone.

**Results:** 201 patients were identified. Of which 151 patients received CTX in addition to ST (mean age ± SD 59 ± 13 years, 81% male and mean BMI ± SD 28 ± 2) and 50 patients received ST alone (mean age ± SD 63 ± 12, 64% male and mean BMI ± SD 27 ± 2). We observed that the patients with critical COVID-19 receiving CTX in addition to ST had significantly better outcomes including reduced in-patient mortality (13% versus 40%, p <0.001), length of hospital and critical care unit stay (mean, 11 versus 15 days (p <0.001) and 6 versus 11 days (p <0.001) respectively), and the need for mechanical ventilation (16% versus 42%, p <0.001) with improved CRP at day 7 (mean, 38mg/L versus 62mg/L, p = 0.001).

**Conclusion:** These results may be due to the antibiotic and anti-cytokine effects of CTX. Clinical trials are currently underway to test our observations.

## Introduction

Over 10 million people in India have had confirmed Coronavirus Disease 19 (COVID-19), resulting in >150,000 deaths. The disease is self-limiting for the majority of those infected, but for those with critical disease steroids and supportive measures remains the mainstay of treatment.^1^ Severity of the disease is related to various factors including male sex, obesity, ethnicity, diabetes and prior cardiac or respiratory diseases.2

A proportion of patients with severe COVID-19 mount an exaggerated immune response and react by producing an excess of cytokines.3 T-lymphocyte, monocyte and neutrophil activation as a result of this ‘cytokine storm’ leads to respiratory failure and acute respiratory distress syndrome (ARDS) requiring oxygen therapy and ventilatory support and is associated with higher mortality.4,5

Viral pneumonias are known to predispose to secondary bacterial infections. Staphylococcus Aureus, MRSA and Sternotrophomonous maltophilia are the bacteria commonly seen in patients with Influenza A and in ventilated patients with SARS.^6,7^

Co-trimoxazole (1:5, trimethoprim: sulphamethoxazole) is a bactericidal antibiotic used in the treatment of various infections.^8^ Methicillin sensitive Staphylococcus Aureus (MSSA), Methicillin resistant Staphylococcus Aureus (MRSA), Klebsiella pneumoniae, Haemophilus influenzae B and Stenotrophomonas maltophilia are some of the bacteria it is effective against.^9^

Cotrimoxazole also has effects upon the immune system, although not widely recognised.^10^ The cytokine storm may occur as a result of neutrophil recruitment to the lung due to the stimulation of the formyl peptide receptors (FPRs) by formyl peptides also known as Damage associated Molecular patterns which are released upon the mitochondrial injury of the host cells. FPRs are situated on the outside of the neutrophils and monocytes, and when stimulated cause the release of intracellular and extracellular reactive oxygen series (ROS) which can drive the cytokine activation. In addition it stimulates the formation of Neutrophil Extracellular Traps (NETs) which block the alveolar capillary bed leading to significant hypoxaemia. Co-trimoxazole blocks the FPRs and can reduce the movement of neutrophils to the lung and NETosis. In addition sulphamethaxazole inhibits neutrophil activation by inhibiting Phorbol 12-myristate 13-acetate (PMA) stimulation thereby preventing Protein Kinase C activation and reducing inflammation.^11,12^ This offers a possible explanation for the observed clinical benefit by reducing neutrophil, monocyte and lymphocyte activation leading to a reduction in the risk of ARDS.^12-17^

Case reports describing clinical recovery from acute respiratory distress sydrome after the addition of cotrimoxazole are seen in medical literature.^18^ Recently more data has been published on the benefits of CTX in severe COVID-19.^19-22^

We have treated nearly 700 patients with severe to critical COVID-19 with co-trimoxazole. Here we report our preliminary findings with the use of cotrimoxazole in the treatment of critical COVID-19, compared with a group of concurrent controls.

## Methods

We retrospectively analysed data obtained from electronic case records and case notes of patients admitted to the critical care facility of the IQ City Medical College Hospital, Durgapur, West Bengal, India from 15^th^ June 2020 to 15^th^ November 2020 and followed up until discharge, death or the census date of 30^th^ November 2020. Data was collected, anonymized and stored in a highly secure, password protected database.

Patients admitted with clinically suspected or confirmed COVID-19 were commenced on standard therapy including antibiotics (piperacillin/tazobactam and clarithromycin or azithromycin), dexamethasone, low molecular weight heparin, paracetamol, supplemental oxygen therapy, intravenous fluids, antiviral drugs and other investigational therapies based on ICMR guidelines.^23^ All the patients had Chest X rays and CT chest scan done on admission which confirmed lung infiltrates in a pattern consistent with a radiological diagnosis of COVID-19. All patients fitted the WHO criteria for critical COVID-19.^24^

Cotrimoxazole is licensed for the treatment of respiratory infections and pneumonia. Patients diagnosed with COVID-19 (RT PCR or clinical) with increasing oxygen demands requiring non-invasive ventilation in the form of Continuous Positive Airway Pressure (CPAP) or High Flow Nasal Cannula (HFNC) were administered oral Cotrimoxazole (CTX; 160mg of trimethoprim and 800mg sulphamethoxazole) 8 hrly for 7 to 10 days in addition to standard therapy.^8^ This was agreed and approved by the physicians at the local respiratory departmental meeting held on the 15^th^ of June 2020.

Consecutive cases of newly diagnosed critical COVID-19 receiving Cotrimoxazole in addition to standard therapy were included in the analysis. This group was matched 3:1 with concurrent controls (n=50) for age and disease severity (critical COVID-19).

Mortality, progression to mechanical ventilation, length of stay, improvements in CRP and Neutrophil-Lymphocyte Ratio (NLR) at Day 7 were the outcomes analysed.^25,26^

Day 0 in the cases and controls was the day of admission to hospital.

Data on comorbidities previously suggested to be linked to higher risk of mortality in COVID-19 were also compared between the two groups.

### Statistical Analysis

Continuous data are presented using mean and standard deviation. Comparisons between two groups of continuous data was made using the t-test for parametric and Mann Whitney U-test/Wilcoxon signed rank test for non-parametric data.

Categorical data was presented as number or percentage of patients. For categorical variables comparison between two groups was done using Chi-squared test or Fishers exact test (for small numbers). Prognostic variables were assessed using univariate and multivariate Cox regression analysis for survival. 27 variables in the whole cohort, case and control group were identified based on previous literature and entered into univariate Cox regression analysis.^2^ Variables with a p-value of <0.20 at univariate analysis and < 10% missing data were included in the multivariate analysis using forward logistic regression method. A p-value of < 0.05 was considered to be significant. The statistical software SPSS was used for the analysis.

### Ethics

Ethical approval was obtained from the IQ City Medical College Hospital Institutional Ethics Committee.

## Results

### Baseline charecteristics

Baseline characteristics from anonymized record reviews are shown in table 1 for standard therapy patients and those treated with the addition of CTX to standard therapy. Prior to ‘Day 0’ these patients had symptoms for a mean duration of 4 days in the CTX and control group. The groups were comparable for age, ethnic group, BMI, chronic lung disease, ischemic heart disease and Hypertension. Diabetes and chronic kidney disease were lower in the CTX added group at 39% and 2.7% respectively, compared with standard therapy alone at 60% and 18% respectively. Male sex was higher in the cotrimoxazole group at 81% compared to 64% in the standard therapy only group.

**Table 1:** Baseline characteristics of patients with severe COVID-19 receiving Co-trimoxazole (CTX) with standard therapy or standard therapy alone.

|  | **CTX + standard therapy** | **Standard therapy alone** | **p-value** |
| --- | --- | --- | --- |
| **Subjects** | 151 | 50 |  |
| **Age (years), mean ± SD** | 59 ± 13 | 63 ± 12 | 0.085 |
| **Male, %** | 81 | 64 | 0.015 |
| **Body Mass Index** | 28 ± 2 | 27 ± 2 | 0.209 |
| **Comorbidities, %** |  |  |  |
| Any disease | 88 | 88 | 0.988 |
| Diabetes Mellitus | 39 | 60 | 0.010 |
| Hypertension | 60 | 74 | 0.080 |
| Dyslipidemia | 3 | 0 | 0.245 |
| Chronic obstructive pulmonary disease | 11 | 8 | 0.595 |
| Ischemic Heart Disease | 10 | 8 | 0.685 |
| Asthma | 3 | 0 | 0.245 |
| Congestive cardiac failure | 1 | 0 | 0.413 |
| Chronic kidney disease | 3 | 18 | 0.000 |
| Interstitial Lung Disease | 2 | 0 | 0.315 |
| **Symptoms** |  |  |  |
| **Duration of illness (days)** | **4 ± 2** | **4 ± 2** | **0.843** |
| **Breathlessness, %** | **100** | **99** | **0.081** |
| **Fever, %** | **42** | **40** | **0.767** |
| **Cough, %** | **56** | **50** | **0.439** |
| **Chest pain,%** | **6** | **0** | **0.077** |
| **Diagnosis** |  |  |  |
| RT-PCR positive/CT changes, % | 79/21 | 60/40 | <0.001 |
| **Day 0** |  |  |  |
| **Clinical parameters, mean ± SD** |  |  |  |
| Respiratory rate (breaths/min) | 31 ± 5 | 30 ± 4 | 0.137 |
| Body temperature (°F) | 99.3 ± 1.5 | 99.4 ± 1.3 | 0.766 |
| C-Reactive Protein (mg/L) | 98 ± 30 | 98 ± 34 | 0.721 |
| Neutrophil Lymphocyte ratio | 10.6 ± 10.3 | 9.1 ± 5 | 0.604 |
| **Ventilatory support** |  |  |  |
| CPAP / HFNC, % | 87/13 | 85/15 | 0.245 |

Definition of abbreviations: SD = standard deviation; CPAP = Continuous Positive Airway Pressure; HFNC = High Flow Nasal Cannula;
Comparison between continuous variables and categorical variables was made by the Mann-Whitney U test and Chi Square test /Fishers exact test respectively. A p-value of <0.05 was considered statistically significant.

Baseline observations were similar for duration of symptoms, respiratory rate, body temperature, C-reactive protein and NLR (table 1). All patients required either CPAP or HFNC fulfilling the criteria for critical COVID-19.^24^

### Outcome

87% of patients with added CTX were discharged well without oxygen after a mean length of hospital and critical care unit stay of 11 and 6 days respectively, 15% required mechanical ventilation, while 13% succumbed to the illness (table 2).

**Table 2:**
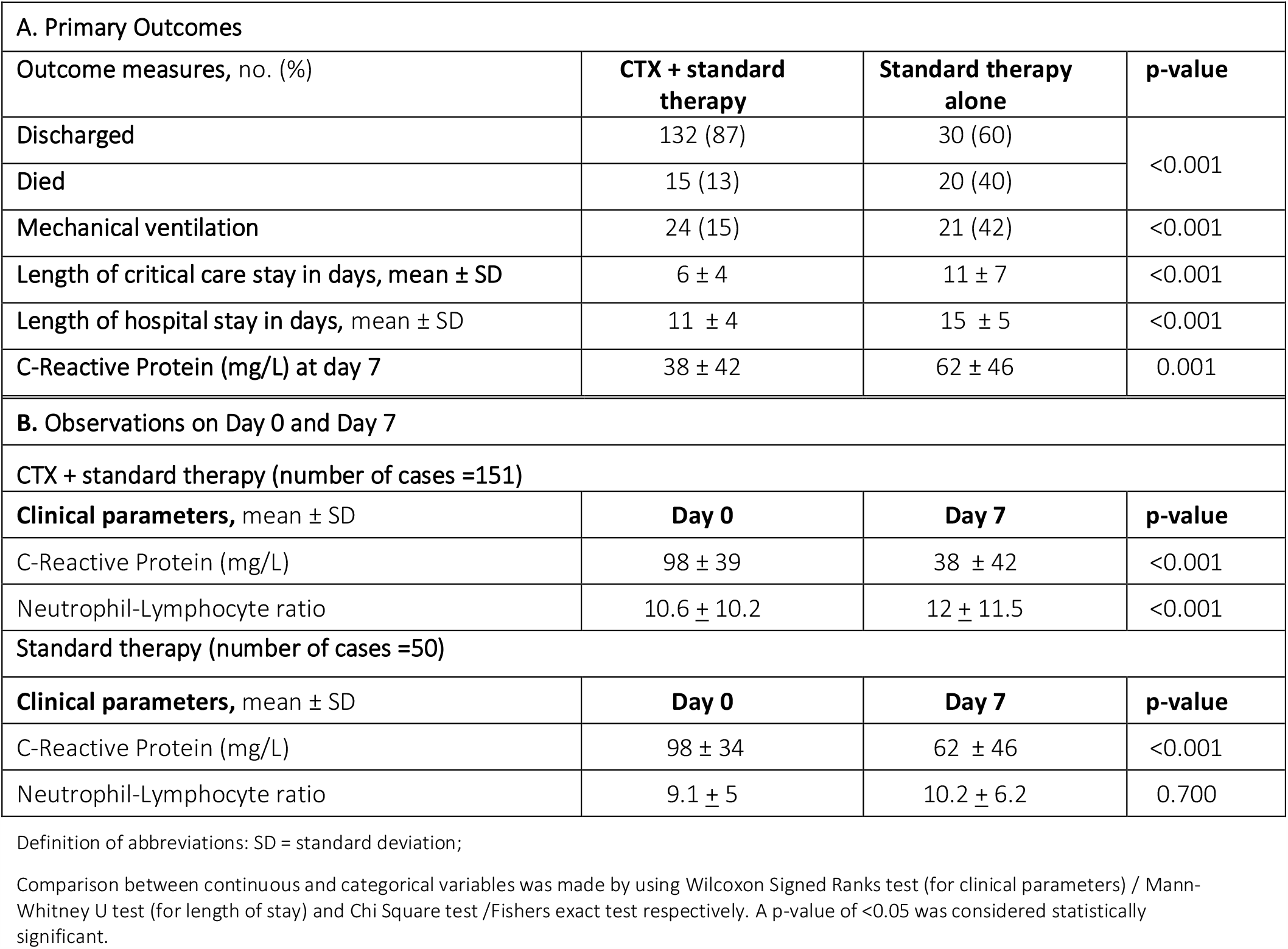
A) Primary outcomes and B) Observations on Day 0 and Day 2 in patients with severe COVID-19 receiving Co-trimoxazole (CTX) with standard therapy or standard therapy alone.

| **A. Primary Outcomes** |  |  |  |
| --- | --- | --- | --- |
| **Outcome measures, no. (%)** | **CTX + standard therapy** | **Standard therapy alone** | **p-value** |
| Discharged | 132 (87) | 30 (60) | <0.001 |
| Died | 15 (13) | 20 (40) |  |
| Mechanical ventilation | 24 (15) | 21 (42) | <0.001 |
| Length of critical care stay in days, mean ± SD | 6 ± 4 | 11 ± 7 | <0.001 |
| Length of hospital stay in days, mean ± SD | 11 ± 4 | 15 ± 5 | <0.001 |
| C-Reactive Protein (mg/L) at day 7 | 38 ± 42 | 62 ± 46 | 0.001 |
| **B. Observations on Day 0 and Day 7** |  |  |  |
| **CTX + standard therapy (number of cases =151)** |  |  |  |
| **Clinical parameters, mean ± SD** | **Day 0** | **Day 7** | **p-value** |
| C-Reactive Protein (mg/L) | 98 ± 39 | 38 ± 42 | <0.001 |
| Neutrophil-Lymphocyte ratio | 10.6 ± 10.2 | 12 ± 11.5 | <0.001 |
| **Standard therapy (number of cases =50)** |  |  |  |
| **Clinical parameters, mean ± SD** | **Day 0** | **Day 7** | **p-value** |
| C-Reactive Protein (mg/L) | 98 ± 34 | 62 ± 46 | <0.001 |
| Neutrophil-Lymphocyte ratio | 9.1 ± 5 | 10.2 ± 6.2 | 0.700 |

Definition of abbreviations: SD = standard deviation;
Comparison between continuous and categorical variables was made by using Wilcoxon Signed Ranks test (for clinical parameters) / Mann-Whitney U test (for length of stay) and Chi Square test /Fishers exact test respectively. A p-value of <0.05 was considered statistically significant.

Data from the patients receiving standard therapy alone, showed that 60% of patients were discharged with a mean length of hospital and critical care unit stay of 15 and 11 days respectively, 42% required mechanical ventilation, while 40% died.

At day 7 patients in both groups showed a significant reduction in C-reactive protein levels compared to day 0, however the reduction was more pronounced in the CTX+ST group compared to the ST only group. In addition, patients with added CTX showed a significant improvement in NLR that was not seen in the group on standard therapy alone. (table 2).

None of the patients in the cotrimoxazole group experienced any side effects warranting stopping of the medication.

### Prognostic indicators

Univariate and multivariate analysis of the whole cohort, cotrimoxazole with ST and standard therapy only groups identified a number of predictors of outcome (supplementary table S1). In the ST only group male sex (p = 0.191), DM (p=1.00) and CKD (p = 0.299) did not predict survival at univariate analysis.

## Discussion

To the best of our knowledge, this is the first study primarily focussing on pharmacological interventions in patients with COVID-19 on non-invasive ventilation. We have compared the baseline characteristics and clinical outcomes in patients who received high dose CTX in addition to ST with patients who received standard only, in a critical care setting.

In this study we have shown a significant reduction in mortality, intubation rate, length of hospital and critical care stay and inflammatory markers in the CTX+ST group compared to the ST only group in keeping with the study by Quadery and co-workers.^19^ The mortality and the rate of intubation in the ST only group was similar to that of current literature where patients with COVID 19 on non-invasive ventilation who were for further escalation was 47% and 57% respectively.^27^

The higher dose cotrimoxazole used in our study is indicated in patients with nocardiasis with excellent safety profile.^28^ This results in higher concentration of the trimethoprim in the blood which is a potentent anti-inflammatory agent resulting in increased inactivation of neutrophils and monocytes in the blood, thus blocking the cytokine storm mediated ARDS.

While male sex, Diabetes & Chronic Kidney Disease were not matched between the CTX+ST and ST only group in this study, a univariate analysis of the cox regression model had not shown any of them to be predicators of mortality in the ST only group suggesting that although these three factors were not matched between the two groups, they did not affect survival in patients receiving standard therapy only.

Cotrimoxazole is an inexpensive drug licensed for use in respiratory infections with few serious side effects. They are generally available worldwide, and may have benefit in preventing acute lung injury in this pandemic.^29^

A randomized control trial with Cotrimoxazole in patients with severe COVID-19 is underway (Clinical Trials Registration-India ID CTRI/2020/10/028297).^30^

### Limitations

Given that this is a case control study, there may be a potential of selection bias. Despite being a retrospective observational study there was excellent data completeness particularly because the patients were closely monitored in a critical care setting. Use of other investigational drugs as part of ST could have been confounding factors, but due to the significantly large difference in the outcomes and no other investigational drug found to be effective in COVID-19 it is obvious that despite the subtle differences in treatment the impact of co-trimoxazole cannot be overlooked.

## Conclusion

If our observation is confirmed by other large scale studies, then hundreds of thousands of lives could be potentially saved worldwide by the use of this inexpensive, widely avalable and safe drug. In addition reducing hospital stay and use of ventilators could potentialy reduce the overwhelming pressure upon the healthcare resources of developing nations. Furthermore this will improve public confidence in the healthcare system and improve and maintain global economy.

## Data Availability

This is a retrospective case control study and all anonymised data collected is stored securely for retrieval if required.

## SUPPLEMENTARY TABLE

**Table S1:**
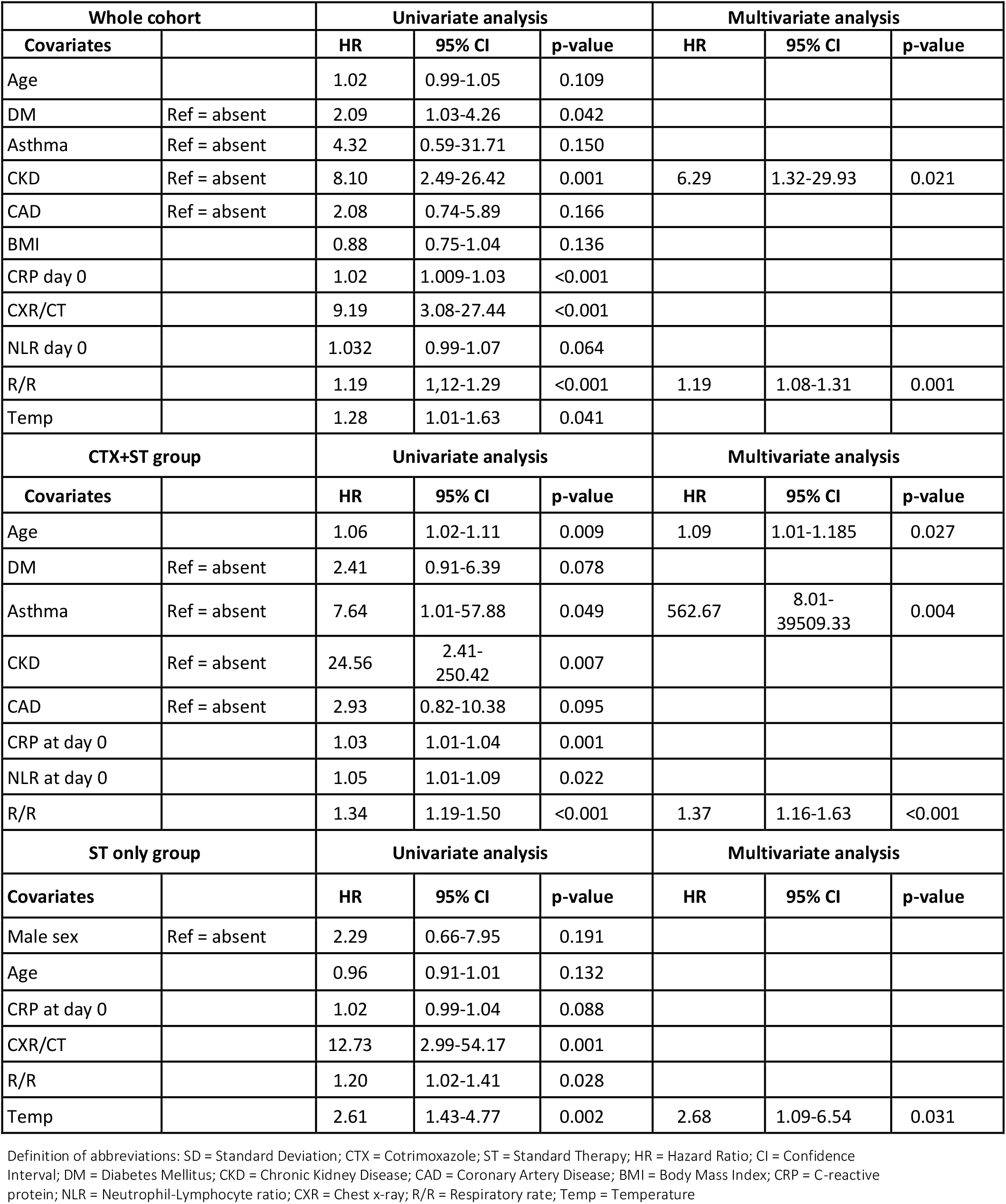
Cox regression survival analysis for the whole cohort, CTX+ST and ST only groups.

## Notes

### Competing Interest Statement

The authors have declared no competing interest.

### Funding Statement

No funding was received

### Author Declarations

IQ City Medical College Hospital Institutional Ethics Committee, Durgapur, West Bengal, India.

