## Supplemental Table 1 for "The impact of high dose oral cotrimoxazole in patients with COVID-19 with hypoxic respiratory failure requiring non-invasive ventilation: A Case Control Study"

### SUPPLEMENTARY TABLE

**Table S1: Cox regression survival analysis for the whole cohort, CTX+ST and ST only groups**

| Whole cohort | | Univariate analysis | | | Multivariate analysis | | |
|---|---|---|---|---|---|---|---|
| **Covariates** | | **HR** | **95% CI** | **p-value** | **HR** | **95% CI** | **p-value** |
| Age | | 1.02 | 0.99-1.05 | 0.109 | | | |
| DM | Ref = absent | 2.09 | 1.03-4.26 | 0.042 | | | |
| Asthma | Ref = absent | 4.32 | 0.59-31.71 | 0.150 | | | |
| CKD | Ref = absent | 8.10 | 2.49-26.42 | 0.001 | 6.29 | 1.32-29.93 | 0.021 |
| CAD | Ref = absent | 2.08 | 0.74-5.89 | 0.166 | | | |
| BMI | | 0.88 | 0.75-1.04 | 0.136 | | | |
| CRP day 0 | | 1.02 | 1.009-1.03 | <0.001 | | | |
| CXR/CT | | 9.19 | 3.08-27.44 | <0.001 | | | |
| NLR day 0 | | 1.032 | 0.99-1.07 | 0.064 | | | |
| R/R | | 1.19 | 1,12-1.29 | <0.001 | 1.19 | 1.08-1.31 | 0.001 |
| Temp | | 1.28 | 1.01-1.63 | 0.041 | | | |
| **CTX+ST group** | | **Univariate analysis** | | | **Multivariate analysis** | | |
| **Covariates** | | **HR** | **95% CI** | **p-value** | **HR** | **95% CI** | **p-value** |
| Age | | 1.06 | 1.02-1.11 | 0.009 | 1.09 | 1.01-1.185 | 0.027 |
| DM | Ref = absent | 2.41 | 0.91-6.39 | 0.078 | | | |
| Asthma | Ref = absent | 7.64 | 1.01-57.88 | 0.049 | 562.67 | 8.01-39509.33 | 0.004 |
| CKD | Ref = absent | 24.56 | 2.41-250.42 | 0.007 | | | |
| CAD | Ref = absent | 2.93 | 0.82-10.38 | 0.095 | | | |
| CRP at day 0 | | 1.03 | 1.01-1.04 | 0.001 | | | |
| NLR at day 0 | | 1.05 | 1.01-1.09 | 0.022 | | | |
| R/R | | 1.34 | 1.19-1.50 | <0.001 | 1.37 | 1.16-1.63 | <0.001 |
| **ST only group** | | **Univariate analysis** | | | **Multivariate analysis** | | |
| **Covariates** | | **HR** | **95% CI** | **p-value** | **HR** | **95% CI** | **p-value** |
| Male sex | Ref = absent | 2.29 | 0.66-7.95 | 0.191 | | | |
| Age | | 0.96 | 0.91-1.01 | 0.132 | | | |
| CRP at day 0 | | 1.02 | 0.99-1.04 | 0.088 | | | |
| CXR/CT | | 12.73 | 2.99-54.17 | 0.001 | | | |
| R/R | | 1.20 | 1.02-1.41 | 0.028 | | | |
| Temp | | 2.61 | 1.43-4.77 | 0.002 | 2.68 | 1.09-6.54 | 0.031 |

Definition of abbreviations: *SD = Standard Deviation; CTX = Cotrimoxazole; ST = Standard Therapy; HR = Hazard Ratio; CI = Confidence Interval; DM = Diabetes Mellitus; CKD = Chronic Kidney Disease; CAD = Coronary Artery Disease; BMI = Body Mass Index; CRP = C-reactive protein; NLR = Neutrophil-Lymphocyte ratio; CXR = Chest x-ray; R/R = Respiratory rate; Temp = Temperature*
